# Problematic Usage of the Internet: Converging and Diverging Terminologies and Constructs

**DOI:** 10.1101/2025.09.08.25335292

**Authors:** Mehran Zare-Bidoky, Alexander M. Baldacchino, Zsolt Demetrovics, Naomi A. Fineberg, Mostafa Kamali, Yasser Khazaal, Marc N. Potenza, Dan J. Stein, Hamed Ekhtiari

**Author notes:** **Corresponding Author:** Hamed Ekhtiari, MD, PhD, Laureate Institute for Brain Research, 6655 S Yale Ave, Tulsa, OK 74136-3326, Department of Psychiatry and Behavioral Sciences, University of Minnesota.

## Abstract

Problematic usage of the internet (PUI) is an increasing global concern, leading to significant negative impacts in various levels. PUI is an umbrella term comprised of subcategories, including online gaming disorder, online gambling disorder, online buying-shopping disorder, online compulsive sexual behavior disorder, problematic usage of social media, cyberchondria, cyberbullying, and digital hoarding. With respect to the diversity of these categories, a factor that further impedes the field of PUI is not recognizing these subcategories under the PUI. To estimate the state of the field, we conducted a systematic review on a sample time frame, 2022-2023, and we found that among 2497 relevant papers, only 13% of the studies on PUI subcategories used PUI as an umbrella term. Establishing PUI as an overarching terminology, as we do with substance use disorder (SUD) as a term while respecting the diversities within its subcategories, would synergize research efforts, facilitate comparisons, and inform treatment strategies. As technology continues to evolve, terminological refinement is critical to maintaining relevance and accessibility for researchers and the broader community.

## Introduction

Problematic usage of the internet (PUI) is an escalating global concern, characterized by engaging in excessive or poorly controlled use of online technologies, including impaired control and shifts in priorities away from important responsibilities, leading to negative familial, social, educational, and occupational consequences (Aboujaoude, 2010; Shaw & Black, 2008). Additionally, PUI has been linked to depression, impulsivity, aggression, social anxiety, attention-deficit hyperactivity disorder (ADHD), and suicidal ideation (Chun et al., 2020; Dang et al., 2024; He et al., 2024; Koncz et al., 2023; Melca et al., 2023; Teng et al., 2021, p. 9). The prevalence of PUI has increased in the past two decades for reasons including growing reliance on the internet in daily life, lockdowns during the COVID-19 pandemic, and the development of addictive design and manipulative algorithms (Burkauskas et al., 2023; Montag et al., 2019; Pan et al., 2020).

Other well-known terminologies, such as “internet addiction,” “compulsive internet use,” “pathological internet overuse,” “internet dependence,” and others, have emerged from varying perspectives, with some focusing more on having clear diagnostic criteria and others formed more around the behavioral mechanisms leading to this phenomenon. The variety of presumptions about the causality and consequences of this phenomenon makes it difficult to reach a coherent concept and subsequent practice. By proposing the term PUI, we take into account this complex phenomenon, without labeling any cause or effect. For instance, the term “internet addiction” is claimed to be overpathologizing and controversial, requiring a subtler term, and the fact that many people use the internet as a means to pathologically engage in other activities, therefore, the internet is not the problem per se (Pezoa-Jares, 2012). The term PUI is an umbrella construct referring to excessive engagement in a variety of online behaviors, including those relating to gaming, gambling, pornography, and social media (Fineberg et al., 2018). For instance, screen-related problematic behaviors involving devices such as smartphones, laptops or tablets fall under the PUI broad category only when the problematic behavior involves excessive engagement with internet-based content, such as social media use and online gaming, rather than with the device itself, as they may be considered a delivery mechanism rather than the focus of an independent disorder. However, there are new emerging forms of PUI like problematic use of AI-chatbot (Yao et al., 2025), problematic online streaming (Singh et al., 2021), and problematic cryptocurrency trading (Jain et al., 2025). When such behaviors are sufficiently problematic to deserve a diagnosis, this may or may not be recognized by existing classification systems. For instance, online gaming disorder (code: 6c51.0) and online gambling disorder (code: 6c50.1) are included in ICD-11 (Brand et al., 2020; Brand & Potenza, 2023; World Health Organization, 2018), whereas behaviors like digital hoarding are at the early stages of scientific recognition (Brand et al., 2020; Sweeten et al., 2018; van Bennekom et al., 2015).

As digital technologies change and scientific knowledge accumulates, the field may benefit from ongoing attention to appropriate terminologies. The European Network for Problematic Usage of the Internet (Fineberg et al., 2022) proposed eight major forms of problematic internet-use behavior to be considered under PUI: (1) online gaming disorder, (2) online gambling disorder, (3) online buying-shopping disorder, (4) online compulsive sexual behavior disorder, (5) problematic usage of social media, (6) cyberchondria (excessive, repeated online searching for illness-related information), (7) cyberbullying (repetitive digital posting of threatening or disparaging messages to another individual), (8) and digital hoarding (excessive accumulation of digital material such as emails, files, media, and software). Given terminological inconsistencies, a factor that may hinder reaching a consensus among scientists is not recognizing these categories of behavioral problems under a general construct, namely PUI.

The case of PUI is in part akin to substance use disorders (SUDs). The concept of an SUD as defined in the DSM-5 provides an umbrella framework that identifies shared characteristics across various SUDs, despite the unique effects of individual substances (American Psychiatric Association, 2013). This approach underscores common features, including impaired control, social impairment, risky use, and other features like tolerance and withdrawal (Ekhtiari et al., 2021), allowing researchers and clinicians to comprehensively address the core aspects of SUDs. This framing helps create a cohesive understanding of addiction as a cross-diagnostic phenomenon, encompassing both unique aspects of individual substances and shared characteristics across substances. On the other hand, the specific nomenclatures for each type of SUD maintain their unique entities. Other conceptual frameworks, such as the obsessive-compulsive and related disorders (OCRDs) model, also utilize the same approach. Diagnostic frameworks such as the DSM-5 and ICD-11 have also adopted an ‘umbrella’ approach, with a wide range of disorders sharing core aspects of compulsivity but with distinct underlying psychobiologies listed within the same OCRD grouping. Indeed, an international case vignette field trial revealed the clustering approach to OCRDs to be helpful for recognition of these disorders by clinicians and produced more accurate diagnoses for many disorders (Kogan et al., 2020). The field of PUI may benefit from employing the same concept. This dual-level approach enables considering common features of disorders under a PUI terminology, while maintaining specific aspects for evaluation, diagnosis, and treatment.

Therefore, we assessed terms used by researchers when conducting studies on PUI in general or on particular subcategories. We conducted a systematic review of the PubMed database using a comprehensive range of general terms such as PUI and more specific subcategories. Our goal was to assess how often researchers use these general and specific terms and how often manuscripts used both a general term and a specific term.

## Methods

### Eligibility criteria

#### Study design and methodology

We were interested in every original paper studying human subjects diagnosed or at risk of problematic usage of the internet (PUI) or its subcategories.

#### Information Source

Studies were identified using the Medline/PubMed (https://pubmed.ncbi.nlm.nih.gov/) database. Relevant articles were identified using the comprehensive search strategy as outlined below. The search results were downloaded in comma-separated format, and the required data, including author information, doi, PMID, article title, and publication year, were entered into an Excel spreadsheet.

#### Language

Only publications with their full text in English were included.

#### Search Strategy

Considering the subject of the review, a list of two sets of keywords was compiled by M.Z.B and M.K. These terms were adapted for use in the PubMed repository, and the exact search syntax and search results are outlined in Table 1. To further validate our search terms, we looked into the available systematic reviews in the field of PUI (https://osf.io/8uc2w/). The first set includes PUI general search terms, and the next 8 sets consist of search terms specific to each subcategory. The date was limited from 1 January 2022 to 31 December 2023.

**Table 1:** A categorical search strategy on problematic usage of the internet (PUI) on PubMed.

|  |  |
| --- | --- |
| General terms | “Internet addiction”[tiab:~0] OR “internet usage”[tiab:~0] OR “internet dependence”[tiab:~0] OR “problematic internet”[tiab:~0] OR “pathological internet”[tiab:~0] OR “online addiction”[tiab:~0] OR “compulsive internet”[tiab:~0] OR “internet overuse”[tiab:~0] OR “Internet disorder”[tiab:~0] OR “Internet Overuse”[tiab:~0] OR “smartphone addiction”[tiab:~0] OR “phone addiction”[tiab:~0] |
| Online Gaming Disorder | (patholog*[Title/Abstract] OR problem*[Title/Abstract] OR addict*[Title/Abstract] OR compulsive[Title/Abstract] OR dependen*[Title/Abstract] OR disorder*[Title/Abstract]) AND (video[Title/Abstract] OR computer[Title/Abstract]) AND (gaming[Title/Abstract] OR game[Title/Abstract]) AND (Internet[Title/Abstract] OR On-line[Title/Abstract] OR Online[Title/Abstract] OR Cyber*[Title/Abstract] OR Digital[Title/Abstract] OR Virtual[Title/Abstract]) OR “internet gaming disorder”[Title/Abstract] OR “video game addiction”[Title/Abstract] |
| Online Gambling Disorder | (Internet[Title/Abstract] OR Online[Title/Abstract] OR Cyber*[Title/Abstract]) AND (gambl*[Title/Abstract] OR gambl*[MeSH]) |
| Online Buying-Shopping Disorder | (Internet[Title/Abstract] OR Online[Title/Abstract] OR Cyber*[Title/Abstract] OR Electronic[Title/Abstract] OR Digital[Title/Abstract]) AND (“buying addiction”[tiab:~0] OR “addictive buying”[tiab:~0] OR “compulsive buying”[tiab:~0] OR “impulsive buying”[tiab:~0] OR “problematic buying”[tiab:~0] OR “pathological buying”[tiab:~0] OR “excessive buying”[tiab:~0] OR “compensatory buying”[tiab:~0] OR “obsessive buying”[tiab:~0] OR “buying disorder”[tiab:~0] OR “shopping addiction”[tiab:~0] OR “addictive shopping”[tiab:~0] OR “compulsive shopping”[tiab:~0] OR “impulsive shopping”[tiab:~0] OR “problematic shopping”[tiab:~0] OR “pathological shopping”[tiab:~0] OR “excessive shopping”[tiab:~0] OR “compensatory shopping”[tiab:~0] OR “obsessive shopping”[tiab:~0] OR “shopping disorder”[tiab:~0] OR “spending addiction”[tiab:~0] OR “addictive spending”[tiab:~0] OR “compulsive spending”[tiab:~0] OR “impulsive spending”[tiab:~0] OR “problematic spending”[tiab:~0] OR “pathological spending”[tiab:~0] OR “excessive spending”[tiab:~0] OR “compensatory spending”[tiab:~0] OR “obsessive spending”[tiab:~0] OR “spending disorder”[tiab:~0] OR “purchasing addiction”[tiab:~0] OR “addictive purchase”[tiab:~0] OR “compulsive purchase”[tiab:~0] OR “impulsive purchase”[tiab:~0] OR “problematic purchase”[tiab:~0] OR “pathological purchase”[tiab:~0] OR “excessive purchase”[tiab:~0] OR “compensatory purchase”[tiab:~0] OR “obsessive purchase”[tiab:~0] OR “purchasing disorder”[tiab:~0] OR “buying problem”[tiab:~0] OR “shopping problem”[tiab:~0] OR “spending |
|  | problem"[tiab:~0] OR "purchasing problem"[tiab:~0] OR shopaholic[Title/Abstract] OR oniomania[Title/Abstract] OR overshoping[Title/Abstract] OR overspending[Title/Abstract]) |
| Cyberchondria | cyberchondria[Title/Abstract] OR ((online[Title/Abstract] OR internet[Title/Abstract] OR web[Title/Abstract]) AND (hypochondr*[Title/Abstract] OR reassur*[Title/Abstract])) |
| Compulsive Sexual Behavior Disorder | "online sex"[tiab:~0] OR sexting[Title/Abstract] OR "online sex disorder"[tiab:~0] OR porn*[Title/Abstract] OR Cyberporn*[Title/Abstract] OR Cybersex*[Title/Abstract] |
| Cyberbullying | (Internet[Title/Abstract] OR Online[Title/Abstract] OR Cyber*[Title/Abstract]) AND (bullying[Title/Abstract] OR aggress*[Title/Abstract]) |
| Problematic Social Media Use | "Social media addiction"[tiab:~0] OR "Social media problematic use"[tiab:~0] OR "problematic Social media use"[tiab:~0] OR "Social media disorder"[tiab:~0] OR "Social media abuse"[tiab:~0] OR "problematic usage of the internet"[tiab:~0] OR "problematic usage of internet"[tiab:~0] OR "Social media misuse"[tiab:~0] OR "Social media compulsive use"[tiab:~0] OR "Compulsive Use of Social Media"[tiab:~0] OR "Excessive Social Media use"[tiab:~0] OR "social media addiction"[tiab:~0] OR "social networking site dependence"[tiab:~0] OR "social media dependence"[tiab:~0] OR "pathological social networking site use"[tiab:~0] OR "pathological social media use"[tiab:~0] OR "compulsive social networking site use"[tiab:~0] OR "compulsive social media use"[tiab:~0] OR "excessive social networking site use"[tiab:~0] OR "excessive social media use"[tiab:~0] |
| Online Digital Hoarding | (Internet[Title/Abstract] OR Online[Title/Abstract] OR Cyber*[Title/Abstract] OR Electronic[Title/Abstract] OR Digital[Title/Abstract] OR Virtual[Title/Abstract]) AND hoard*[Title/Abstract] |

#### Data management

Search results were imported into Excel. Screening of articles for relevance was performed by reviewing the titles and abstracts. All raw and processed data tables were version-tracked, with the latest version available on the OSF repository.

#### Study selection

Screening forms were developed, and studies were checked by two authors (M.Z.B. and M.K). The authors initially checked the eligibility of fifty randomly chosen studies under the supervision of the corresponding author (H.E.), as a calibration exercise to ensure eligibility criteria are applied consistently. Neither of the review authors was blind to the authors or institutions of the studies.

## Results

Published papers over two years (1 January 2022 till 31 December 2023) with relevant terms (4156 results) were screened and extracted by two independent raters, resulting in 2497 manuscripts, which are visualized in Figure 1.

**Figure 1.**
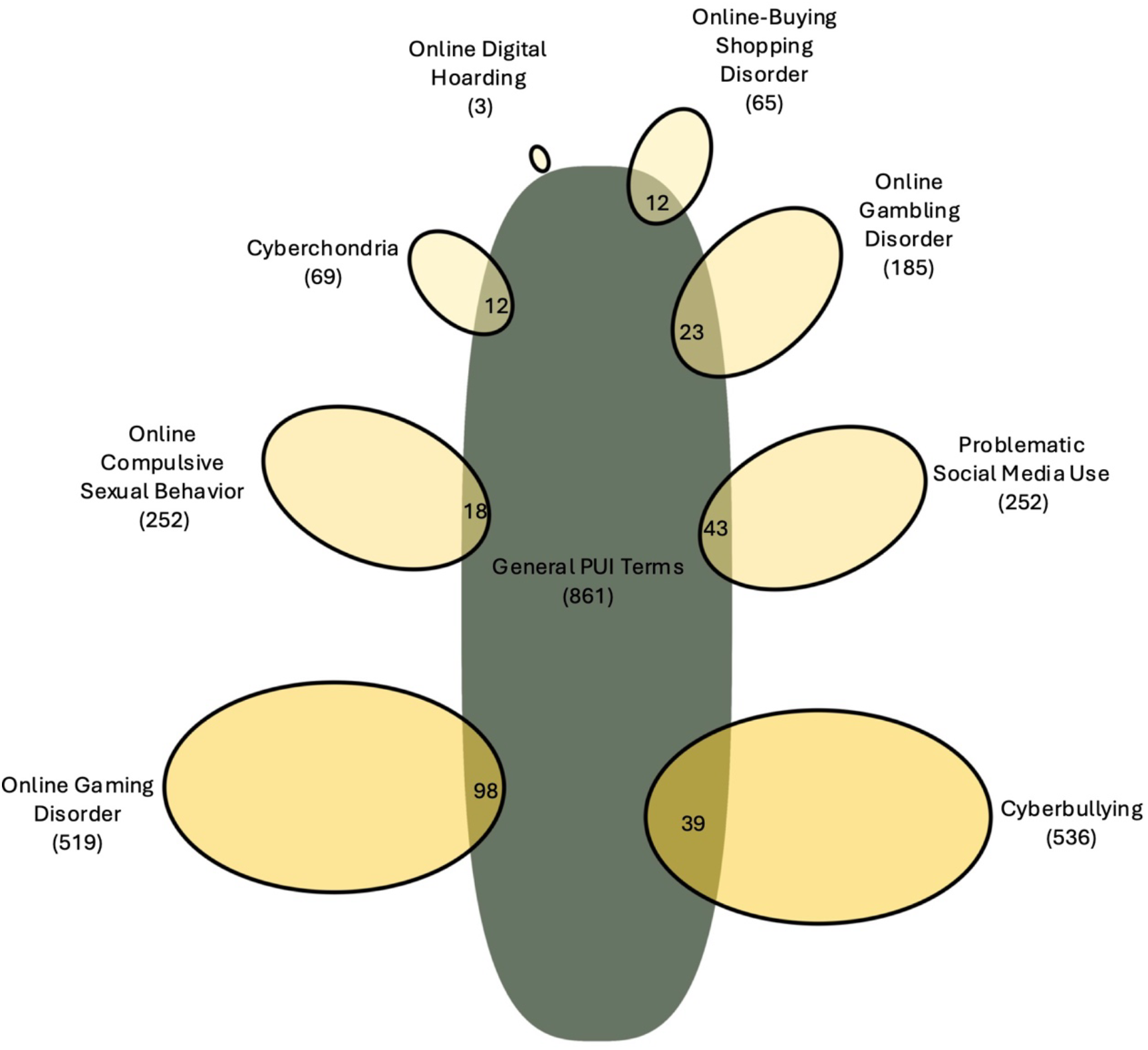
Categories of Online Problematic Behaviors and Their Overlaps with Problematic Usage of the Internet (PUI) based on Terminologies Used in 2108 Published Papers in 2022 and 2023. Overlaps between PUI in general and eight categories of online behaviors: (1) online gaming, (2) online gambling, (3) online buying-shopping disorder, (4) cyberchondria, (5) online compulsive sexual behaviors, (6) cyberbullying, (7) problematic usage of social media, and (8) online digital hoarding are depicted based on whether they only used terminologies related to their specific field or also addressing terminologies related to PUI in general. This figure illustrates the number of included studies retrieved from search terms of problematic usage of the internet (PUI) in general and its subcategories on the PubMed database in the 2022-2023 period. The number of overlapping studies of each subcategory with the general search terms is shown in the intersecting areas.

As shown in Figure 1, the general PUI category comprises the highest number of studies (861), followed by cyberbullying (536), compulsive sexual behavior disorder (252), problematic usage of social media (252), and online gaming (519). Subcategories like cyberchondria (69) and digital hoarding (3) are among the least investigated subjects. Some studies (245) addressed both a general term and a specific category, with the most common overlaps being online buying-shopping disorder (18.4%) and problematic usage of social media (18.1%).

## Discussion

Our findings indicate that, on average, only 13% of articles in the PUI subcategories used general PUI terms in their titles or abstracts (Figure 1). Instead, researchers often focused on the specific behavior in question, such as online gaming or social media overuse, without considering the larger context of PUI as a general construct. There are several possible reasons for this. First, where a particular disorder is well recognized, it is more likely that a specific term will be used. Second, research focused on 1 or 2 behaviors may be more likely to use specific terms, while research focused on many behaviors may use more general terms. Third, clinical research is typically not transdiagnostic and so may be more likely to use specific terms, while public health research often cuts across different behaviors and so may be more likely to use general terms. Fourth, limited integration may reflect the early stage and dynamic nature of the field, with ongoing work on conceptualizing and categorizing behaviors, prior to the establishment of broadly accepted constructs and terminologies.

On the one hand, it is appealing to have consistent terminology. On the other hand, different terms may be useful for different purposes. Psychiatry has appropriately been criticized for reifying mental disorders, meaning treating something abstract and complex as if it were a concrete, physical thing (Hyman, 2010). Instead, it has been argued that clinicians should hold diagnoses lightly (Aftab et al., 2024). As technology changes and science grows, further changes in terminology will likely occur. When studying any particular disorder, there are, however, many advantages in agreeing on common measures (Farber et al., 2023). A lack of consensus on such measures may create challenges in comparing studies, assessing prevalence estimates, conducting systematic reviews, and developing treatments that address PUI and its subtypes.

## Conclusion

Taken together, to advance research and improve the quality of the studies, we support the wider use of PUI as a construct that connects multiple compulsive-impulsive-addictive behaviors that have broadly similar phenotypes, core mechanistic features, and shared intervention implications. Alternative but similar constructs, such as “problematic screen use”, which reflect changes in technology, should also be considered. Establishing a coherent nomenclature may increase consistency across studies, facilitate treatment development, and enhance the progress of research in this important growing field. While alternate frameworks or terms might be considered for PUI, PUI is a widely used term, and we have chosen to use it rather than introduce a new term that may have more limited recognition or consensus. We conclude that the use of an umbrella term such as PUI that captures the full extent of the problem has considerable value at the level of research and clinical practice, particularly while we are still delineating specific constituent disorders and conditions. While the adoption of an umbrella term like PUI may risk obscuring disorder-specific features or pathologizing normative behaviors, we believe that careful subcategorization and contextual precision can safeguard specificity while promoting clarity across research and clinical domains. As technology is advancing so rapidly, it is also critically important to ensure that our terminology continues to remain up-to-date, accurate, and easily understood by scientists and the public alike.

## Data Availability

All data produced in the present work are contained in the manuscript

## Authors’ Contribution

All authors contributed to the conceiving of the presented idea and designing the study. MZB, MK, and HE gathered the data, designed the tables, and performed the initial analytical calculations. All authors discussed the results and contributed to the final manuscript

## Acknowledgment

The authors thank M Ebrahimi for contributing to the figure’s design.

